# Physical activity-correlated changes in plasma enzyme concentrations in fragile sarcolemmal muscular dystrophies

**DOI:** 10.1101/2022.04.01.22273213

**Authors:** Paul S. Blank, Adriana E. Golding, Ivonne Morales Benavides, Hang Waters, Elena Mekhedov, Ludmila Bezrukov, Rebecca D. Wachter, Irina Mikhailenko, Robert H. Brown, Carsten G. Bönnemann, Andrew P. Demidowich, Minal S. Jain, Jack A. Yanovski, Joshua Zimmerberg

## Abstract

**Background and Objectives:** Muscular dystrophies associated with decreased sarcolemma integrity lack validated clinical measures of sarcolemma fragility that can be used to assess disease progression and the effects of therapies designed to reduce sarcolemma fragility. We conducted a pilot study to test the hypothesis that physical activity leads to significant changes in muscle-derived plasma enzymes in participants with “fragile sarcolemmal muscular dystrophies” (FSMD).

**Methods:** We enrolled ambulatory individuals clinically affected with genetically confirmed FSMD neither taking anti-inflammatory medications nor having relevant co-morbidities for an inpatient study. Over five days, blood samples at 20 time points were obtained. Plasma enzymes alanine and aspartate aminotransferase (ALT, AST), creatine kinase (CK), and lactate dehydrogenase (LDH), all found in muscle, were measured before and after routine morning activities and motor function testing. Analysis of Z-transformed time series data led to feature and kinetic models that revealed activity-dependent feature and kinetic parameters.

**Results:** Among the 11 enrolled participants, (LGMD Type 2B/R2 Dysferlin-related (4F/1M), LGMD Type 2L/R12 Anoctamin-5-related (3F/2M), LGMD Type 2I/R9 FKRP-related (1M)), plasma enzymes increased with activity. The average % change +/- SEM with morning activity across all participants was ALT 12.8 ± 2.8%, AST 11.6 ± 2.9%, CK 12.9 ± 2.8%, and LDH 12.2 ± 3.9%, suggesting the increases originate from the same stimulated source, presumably skeletal muscle. For ALT, AST, CK, and LDH, characteristic kinetic features include (a) elevated enzyme activities on arrival that decreased overnight; (b) a longer decay trend observed over the week, and (c) for ALT, AST, and CK, a similar decay trend observed with post-morning activity blood draws.

**Discussion:** Controlled activity-dependent changes in plasma ALT, AST, and CK on time scales of days to weeks can serve as common outcome measures for sarcolemma integrity and may be efficient and effective tools for monitoring disease progression and treatment efficacy for both individuals and patient populations. In addition, this study provides data that may benefit patient management as it can inform guidance on duration and type of activity that minimizes muscle damage.

## INTRODUCTION

The sarcolemma experiences repeated and transient injuries by the normal stresses associated with physical activity ^1^. In healthy individuals, the sarcolemma is repaired. However, individuals with a fragile sarcolemma muscular dystrophy (FSMD), caused by function-altering gene variants encoding membrane and repair proteins, leave the sarcolemma prone to excessive damage and impaired healing ^2-4^. Gene variants causing FSMD include abnormalities in Dysferlin (LGMD R2 DYSF-related and Myoshi myopathy), Anoctamin-5 (LGMD R12 ANO-5), and Fukutin-related protein (LGMD R9 FKRP-related). Dysferlin, a calcium-mediated, membrane-associated protein, Anoctamin-5 (ANO5), a putative calcium-activated chloride channel, and Fukutin-related protein (FKRP), a putative glycosyltransferase involved in alpha-dystroglycan glycosylation, are all implicated in membrane repair ^5-12^.

Damage to and subsequent necrosis of the muscle fiber allows release of muscle proteins into the extracellular environment ^1, 2^. In the absence of inflammation, muscle-derived proteins move through interstitial fluid into the lymphatic system, where, as components of the lymph, these proteins are returned to blood. This knowledge is applied in the clinic when diagnosing muscular dystrophy: muscle weakness, in conjunction with abnormally high muscle-derived creatine kinase concentration ([CK]) in plasma or serum, is the primary evidence of sarcolemma fragility ^13-15^. However, although elevated plasma CK concentrations in a young person with a muscular dystrophy phenotype may be confirmatory, a low [CK] requires further diagnostic review. Importantly, due to significant personal and population variability in plasma [CK], a formal diagnosis of FSMD must be genetically confirmed.

Beyond diagnosis, plasma CK is routinely measured at clinic visits, the interpretation of [CK] is limited. There are currently few quantifiable measures of disease severity and progression in patients with FSMD. Activity measures for patients with upper and/or lower limb impairments such as the six-minute walk test (6MWT), ACTIVLIM, and North Star ambulatory assessment (NSAA) serve as common outcome measures for monitoring disease severity and progression in individual and patient populations ^16-18^. These assessments are not ideal because they are not entirely objective: patients’ ability to complete physical tasks varies between individuals and between visits, and for ACTIVLM, physical limitations are self-reported. Quantifiable outcome measures, commonly referred to as “biomarkers” ^19^, are frequently used for monitoring disease severity and progression. A plasma biomarker for membrane fragility would be beneficial for both clinical research and medical practice ^14, 20, 21^. Such a biomarker would permit i) determining the efficacy of therapeutic interventions, ii) defining clinical guidelines for minimizing muscle wasting, and iii) elucidating disease mechanisms to help identify drug targets and therapeutics. A key feature of FSMDs is that these diseases all progress slowly, making traditional trial outcomes, such as decreasing the rate of progression, challenging. Methodologies that facilitate a more rapid identification of treatments that attenuate exertional fragility are needed.

Here, we report findings from a pilot study to assess biomarkers of changes in barrier function of skeletal muscle in individuals with FSMD. We hypothesized that there would be significant, activity-dependent changes in plasma proteins on short timescales (minutes to days) in individuals with FSMD, and that these proteins could serve as biomarkers for sarcolemma fragility. To test this hypothesis, a short-term, time-dependent investigation with multiple blood draws before and after controlled levels of physical activity was conducted. We found significant activity-dependent changes in plasma alanine and aspartate aminotransferase (ALT, AST), CK, and lactate dehydrogenase (LDH). Furthermore, time-series analyses identified three of the four enzymes (ALT, AST, CK) as having similar and reproducible rates of clearance. We propose that time-series analyses of plasma ALT, AST, and CK before and after activity may serve as sensitive and quantifiable biomarkers of sarcolemma fragility.

## MATERIALS AND METHODS

### Standard Protocol Approvals, Registrations, and Patient Consents

An ongoing pilot study (ClinicalTrials.gov Identifier NCT01851447) to assess changes in blood chemistries before and after activity, beginning October 2014, enrolled ambulatory individuals who were clinically affected with genetically confirmed FSMD. Participants were recruited through the Jain Foundation, Muscular Dystrophy Association, and patient/parent support groups. Individuals taking anti-inflammatory medications or having other co-morbidities were excluded as these could be study confounders. The protocol was approved by the NIH Clinical Center and NICHD Institutional Review Board and all participants provided written informed consent for their participation. Data in this report were collected from October 2014 to November 2017.

### Study timeline

All participants were admitted to the NIH Clinical Center for a 5-day inpatient stay. Upon admission, an intravenous (IV) catheter was placed in the arm, and all subsequent blood draws were obtained from this IV or a replacement. Tourniquets were not used to assist with blood draws as they could falsely affect measured enzyme concentrations. On the morning of Day 1, participant health was assessed with baseline labs: Complete Blood Count (CBC), Prothrombin Time (PT), Partial Thromboplastin Time (PTT), Creatinine level, Basic Metabolic Panel, Hepatic Panel, Cystatin C, high-sensitivity C-reactive protein (hsCRP), Hemoglobin A1C, Thyroid Stimulating Hormone, and Free T4 were measured using standard procedures by the Clinical Center Department of Laboratory Medicine.

A total of 20 blood samples were obtained over the 5-day inpatient period (Figure 1): serial blood draws were performed according to the following schedule. Day 1: beginning of bedrest at night; Day 2: 3 AM in supine position, 7 AM after bedrest but prior to any activity in supine position, 30 min following routine morning activity (e.g., toilet, shower, dressing, breakfast), immediately after physical therapy assessment and every 5 min thereafter for 30 min, and 2, 4, 6 hrs post-assessment; Days 3-5: 7 AM before routine morning activity and 30 min following routine morning activity. ALT, AST, CK, and LDH were measured using Roche Cobas analyzers with lower limits of analytical measurement range of 5, 5, 10, and 7 U/L and imprecision (Total CV; %) less than or equal to 3.3, 2.3, 2.7 and 3.3, respectively.

**Figure 1:**
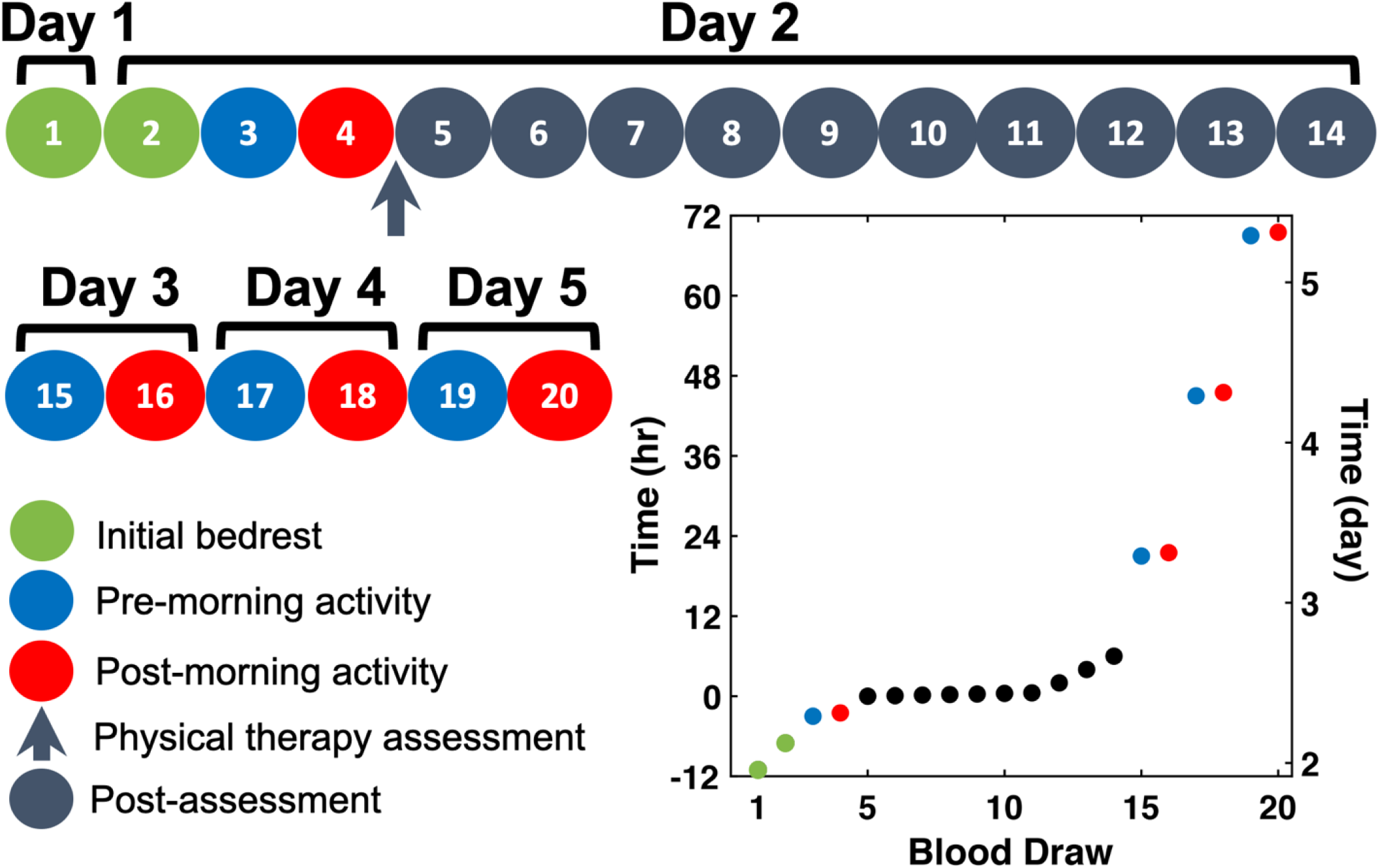
Phlebotomy protocol. Schematic of blood draw schedule over the course of 5 days. Blood was drawn at (1) beginning of bedrest on Day 1, (2) during bedrest on Day 2 at 3AM (green circles); (3, 15, 17, 19) 7AM after bedrest but prior to any activity in supine position (blue circles); (4, 16, 18, 20) 30 min after routine morning activity each day (red circles); (5-14) at high frequency over a short time interval following physical therapy assessment (black circles; time 0 on y-axis, every 5 min for draws 6-11, 90 min between draws 11 and 12, and 120 min between draws 12, 13 and 14).

### Data analysis

Large population variability precludes simple averaging strategies. The enzyme activities were not normally distributed, bounded by the limits of detection, and best described, over all participants, as lognormally distributed. Consequently, all activities were log transformed prior to subsequent data analysis. For group comparisons (t-Test or ANOVA), variance homogeneity was evaluated using Levene’s statistic (absolute) and when heteroscedastic, Welch’s tests were used for group comparisons. Additionally, an exponential process typically used in compartmental analysis, is represented as one or more straight lines modeled using piece-wise continuous lines. Two individuals with the same exponential process but different amplitudes have the same slope for their individually log transformed data. In theory, multiple individuals could be compared through the weighted average of their individual parameters sets. However, more complex models were ill-determined with the limited, time series data available. Therefore, even with log transformation, variability due to differences in individual participant concentrations was reduced by individual Z-score transformation where Z(t_i_) = (Value(t_i_)- μ)/ σ and i = 1-20 (μ = individual’s average and σ = standard deviation (SD) over all draws). Z-score transformation facilitates comparisons of temporal features and structures between sequences with different amplitudes; the average of individual Z-scores therefore describe the behavior of the entire participant cohort. The slope of the log transformed data can be recovered, on average, by dividing the slopes in the Z-score model by the average SD of the cohort data set provided that this parameter is well defined. This was the case for our data. The decay times, appropriate for time dependent compartment models were derived using this approach. Note, the Z-score averages are no longer necessarily Z-distributed but can be renormalized to create a new Z distributed parameter.

Two models describe the Z-score averages. The first, designated a feature model, captured defined data set characteristics using global fitting and constraints on specific time series components. The second, designated a consensus, continuous time series model, used the decay constants derived from the feature model to describe the expected time dependence of a continuous time series. This model may be useful in developing experimental sampling strategies based on the specific features of interest. Both models are formally described in the eMethods.

A consensus time series was created by averaging ALT, AST, and CK data for all 11 participants, with subsequent correlation analysis to remove poorly correlated participant data ^22^. Four individual time series of the original 33 were dropped and a new average and variance were calculated from the remaining 29 time series. This average was fit by the continuous time series model with fixed decay times and established by the feature model, where amplitudes (2 parameters – Amp1 and Amp2), decay fractions in the high frequency region (1 parameter-f1) and ΔZ-score (1 parameter) were evaluated by the fitting (done with weighting using normalized inverse variance).

The expected data set is 220 per analyte, or 880 total. However, a total of 2 blood draws were not collected (8 missing values) and 3 outliers (identified using Matlab outlier detection) were dropped leading to 869 data points analyzed. Data analysis was conducted using Excel and Matlab. Errors are reported as 95% confidence intervals or standard error of the mean (SEM). Figures were created using the data visualization toolbox Gramm (https://doi.org/10.21105/joss.00568).

### Data Availability

Anonymized data not published within this article is available by request from the corresponding author.

## RESULTS

A total of 11 participants were enrolled. The cohort consisted of 5 LGMD Type 2B/R2 Dysferlin-related (4F/1M), 5 LGMD Type 2L/R12 Anoctamin-5-related (3F/2M), and one with LGMD Type 2I/ R9 FKRP-related (1M). All participants exhibited normal kidney function (eGFR range 72 to >120 mL/min/1.73 m^2^). Participants with LGMD 2B were younger than those with LGMD 2L (36.7 +/- 5.3 and 54.1 +/- 1.6; mean +/- SEM, n=5, p = 0.03 unequal variance t-test). There were no differences in the average height and average BMI of the R2 and R12 participants. Participants self-identified Race (64% White, 18% African American, 9% Asian, and 9% Multiracial) and ethnicity (91% Not Hispanic or Latino, 9% Hispanic or Latino).

Plasma ALT, AST, CK, and LDH enzyme activities (concentrations) varied between participants and between LGMD type (R2, R9, R12) (Figure 2A, eFigure 1). Following log transformation, when grouped separately, or together (R2, R12, R2 and R12, and R2, R9, and R12), only ALT (all groups), AST (R2 only), and LDH (R2 only) showed equal variance. ALT, AST, CK, and LDH were lower in R12 compared to R2, over all samples (mean(log(Analyte_R12_))-mean(log(Analyte_R2_)) = -1.01 (−1.14, -0.90), -0.59 (−0.70, -0.50), - 1.62 (−1.78, -1.47), -0.58 (−0.67, -0.50), respectively; mean differences (99% lower, upper bootstrap CI, 10^6^ resamples). Mean values within each group were unequal (ANOVA analysis; p<10^−10^). Consequently, for each analyte, direct averaging across all participants resulted in large uncertainty (95% bootstrap confidence interval; 5,000 resamples) around the average (Figure 2B). Large uncertainties exist even when averaging by LGMD type. The differences between participants (variance and mean) within groups and between groups (inter- and intra-personal variation) highlights the data complexity and the requirement for feature preserving analysis. When each participant’s time course for any one analyte was examined (eFigure 1), similar responses for each analyte were found: decreasing enzyme levels with rest and increasing enzyme levels with activity. The percentage increases of each enzyme with activity were similar, as described below. The large variations between individuals were addressed to both examine correlations between LGMD R2 and R12 populations and the population statistics using Z-score transformation. First, each participants’ log transformed data underwent a Z-score transformation, whereby we report the number of SDs each log data value is from the participant’s own average over all 20 blood draws (eFigure 1). This Z-score transformation was then averaged (Z-score_avg_) over all participants for each blood draw (Figure 2C). The averaging of the Z-transformed data resulted in reduced uncertainty (95% bootstrap confidence interval; 5,000 resamples) around the average. The trends observed in the Z-score_avg_ reflect those of each individual participants’ Z-scores over the course of the visit (Figure 2, eFigure 1). This trend is also seen when plotting the percent change around the average in plasma enzyme levels by blood draw (eFigure 2A).

**Figure 2:**
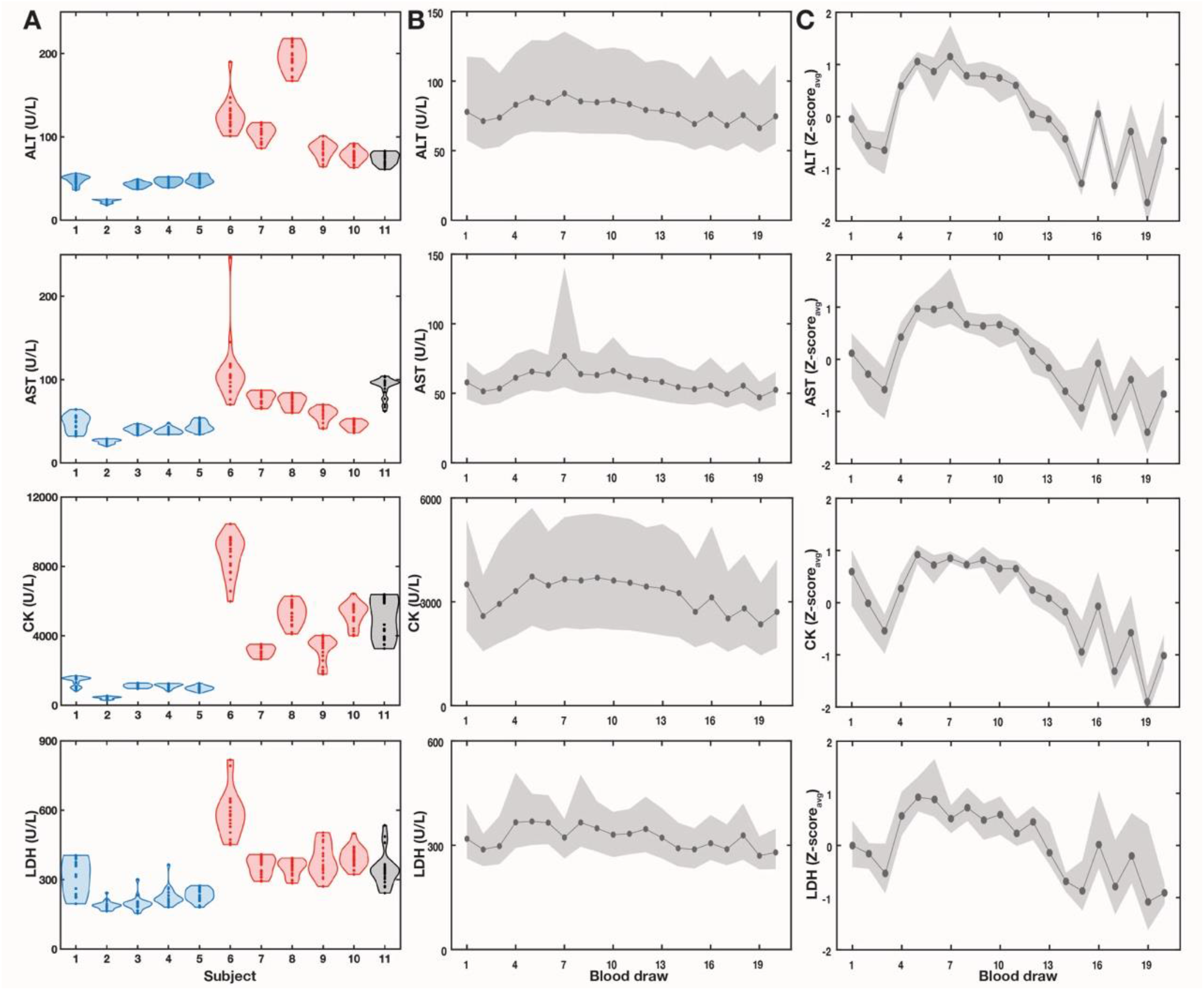
Significant variability in plasma enzymes between participants requires meaningful transformation for time-series analysis of entire cohort averages. A) Violin plots of plasma ALT, AST, CK, and LDH by participant, clustered by LGMD subtype (blue-R12 Anoctamin5-related; red-R2 Dysferlin-related; black-R9 FKRP-related). B) Average over all participants (solid line) and 95% bootstrap confidence interval (shading) for each enzyme by blood draw. C) Average over all participants (solid line) and 95% bootstrap confidence interval (shading) of Z-transformed plasma enzyme by blood draw. The Z-score_avg_ represents the average of all participants’ Z_i_ values calculated using a participant’s plasma enzyme average (μ) and SD (σ), where Z = (Draw_i_ - μ)/ σ; i = blood draws 1-20, and Z_avg_ = <Z_i_>_j_ where j = participants 1-11).

The Z-score_avg_ for each enzyme as a function of time have similar temporal features (Figure 3A). The magnitudes of the activity correlated increases were approximately the same for all enzymes. The LDH Z-score_avg_ 95% confidence intervals are greater than the other enzymes, which is consistent with additional differences observed between AST, ALT, CK and LDH described below.

**Figure 3:**
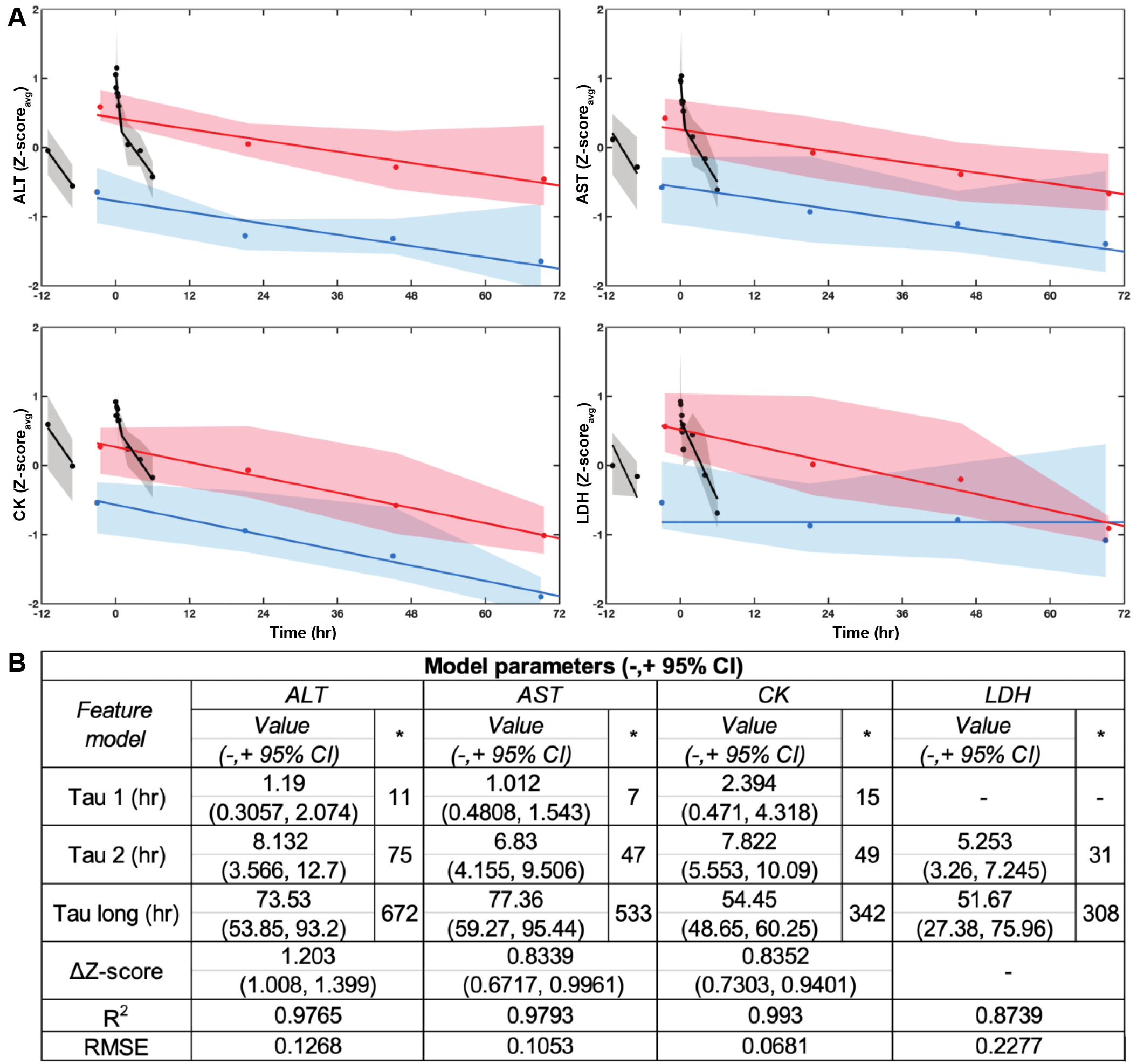
Average plasma enzyme Z-scores reveal three distinct kinetic trends. A) Average (line) and 95% bootstrap confidence interval (shading) of participant plasma ALT, AST, CK, and LDH Z-scores plotted against time (in hours) with physical assessment at time 0. Blue and red displays correspond to pre- and post-morning activity, respectively. Black displays correspond to post-arrival and post-physical assessment. Blue, red, and black displays are associated with similar decreasing, weeklong trends that are uniformly shifted; black is a relatively faster decay process. B) Model parameters for ALT, AST, and CK fit with a feature model consisting of two parallel linear trends and a two-component mixture in the high frequency data region. *Average standard deviation adjusted time constant; R^2^ is adjusted by the number of fitting parameters; root-mean-squared error (RMSE). LDH fit with a feature model consisting of a constant background, a linear trend, and a single decay in the high frequency data region.

The largest increase in plasma ALT, AST, CK, and LDH occurred following routine physical assessment typical at clinic visits (Figures 2, 3). Smaller increases also followed 30 minutes of routine morning activity. Upon arrival at the Clinical Center, plasma enzymes were elevated with the first values comparable to the post assessment values as indicated by ratios ∼1. The average ratios for ALT, AST, CK, and LDH, are 0.90 (+/- 0.04), 0.89 (+/- 0.06), 0.96 (+/- 0.08), and 0.87 (+/- 0.11), (mean +/- 95% CI, n = 11), respectively; this is likely due to high levels of exertion associated with travel. The gradual decline in plasma enzymes over the following four days may be a result of overall decreased activity while hospitalized, as bedrest and reduced physical activity are correlated with reduced plasma enzyme concentrations ^23^.

Other characteristic features observed in the average plasma ALT, AST, and CK Z-score values include (a) elevated enzyme activities on arrival that decreased overnight; (b) two decay components during the high frequency data collection following assessment; (c) a longer decay trend observed over the week that tracks the supine, pre-morning activity blood draws (samples 3, 15, 17, 19), and (d) a similar decay trend observed with post-morning activity blood draws, offset by a constant at each time point (samples 4, 16, 18, 20). These trends were also reflected in the percent plasma enzyme variations around the average over time (eFigure 2B); the average percent change (+/- SEM) in ALT, AST, CK, and LDH across all participants between pre- and post-morning activity were (12.8 ± 2.8%), AST (11.6 ± 2.9%), CK (12.9 ± 2.8%), and LDH (12.2 ± 3.9%), respectively.

### Analysis of data with a new feature model

The similarities and differences between analytes were then evaluated using global fitting of a feature model (Figure 3) revealing that: (a) The longest decay trends are modeled as parallel offset lines, corresponding to single exponential decay processes modified by a constant offset (ΔZ-score). This offset represents the change in Z-score_avg_ that correlated with morning activity. (b) The high-frequency data are modeled as two piecewise, continuous lines corresponding to two exponential decay processes. (c) The initial, overnight decrease in Z-score was best characterized by the larger of the two exponential decay values, evaluated from the high frequency data and described above in (b). While similar, there were noteworthy features in the LDH Z-score values that differed from the other analytes. LDH Z-scores could not be modeled with the same feature set but required a different parameterization where the offset varied with time leading to a different decay trend and the long, supine, pre-morning activity trend was absent and best described by a constant value. That LDH is best described by a different feature model is an important distinction.

Log transformation of an exponentially decaying function results in a linear function. However, upon averaging the transformed Z-scores across the entire cohort, the resulting decay times were weighted by the SD of each individual participants’ data. The time constants obtained by fitting the averaged transformed Z-score data can be converted, on average, to elapsed time unperturbed by the scaling of the slope. This was done after establishing that the SD distributions have well-defined averages and variances and adjusting the decay constants accordingly (Figure 3B). These decay constants can be used in compartment modeling, and for comparison with reported clearance rates.

The feature model for the log transformed analyte data was simplified as the two high frequency time constants were not resolvable. This illustrates an additional advantage for the Z-score treatment as less noise leads to better resolution of multiple decays. To compare with the log data fits, the two constants (τ_1_ and τ_2_) were averaged to obtain an approximate equivalent decay for the single high frequency decay in the log data fits.

The similarities between the fitting parameters of the feature model for plasma ALT, AST, and CK were consistent with a common underlying temporal model. This conjecture is supported by Pearson correlation analysis between the different analytes which revealed high and significant correlations between analytes (eFigure 3). A consensus time series was then developed using the combined plasma ALT, AST, and CK time series data, following removal of minimally correlated time series (4 of 33 with correlation 95% lower limits less than 0; see methods). The consensus time series is described by a 4-parameter continuous time series model capturing the predicted temporal evolution of an analyte described by the model features (kinetic decay constants) identified for plasma ALT, AST, and CK (Figure 4A).

**Figure 4:**
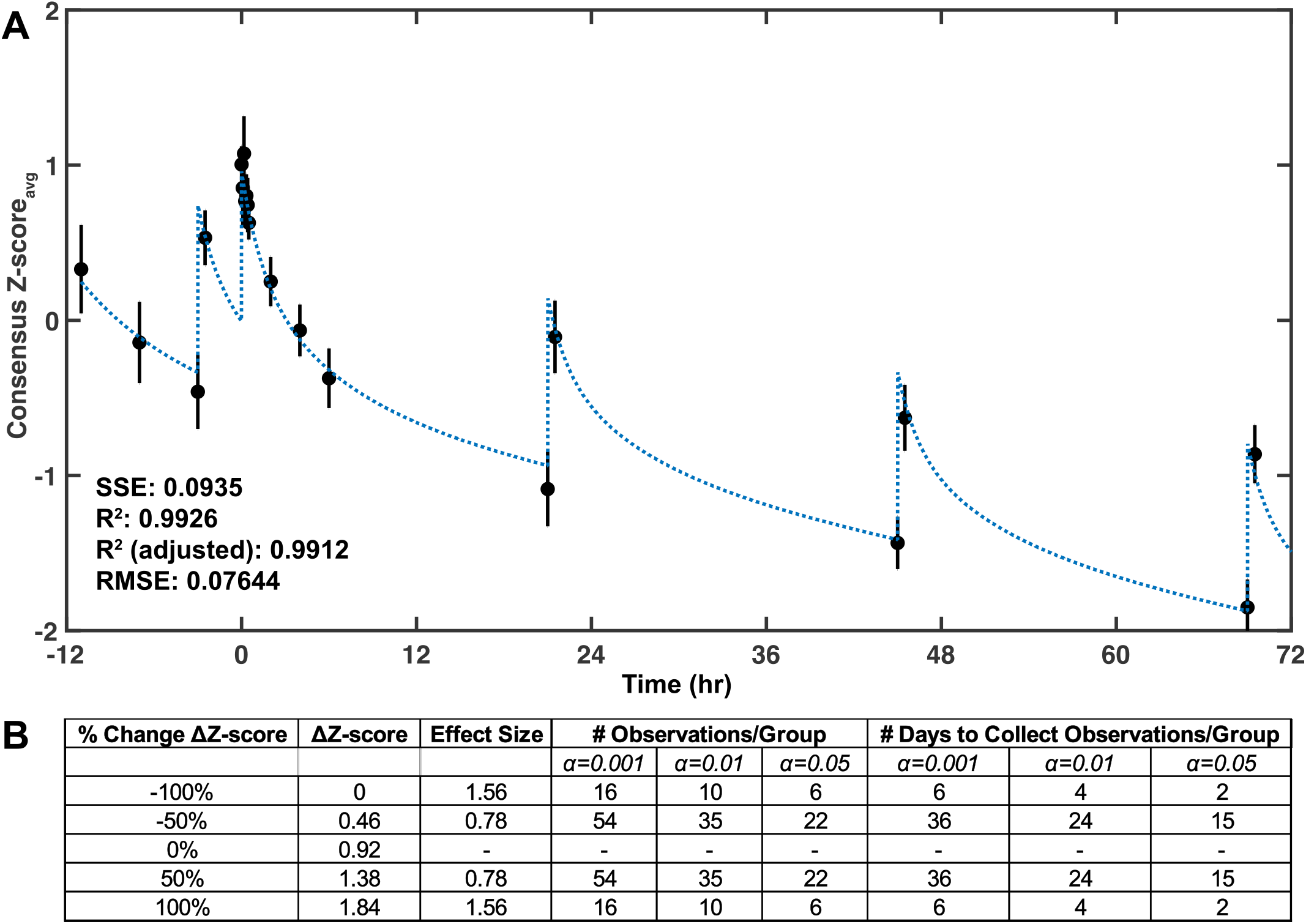
Fitting of cohort consensus data with a continuous time model. A) The data from a consensus dataset including plasma ALT, AST, and CK (but not LDH) was combined and used in developing a continuous time model. The rates determined from the linear slopes in the feature model were fixed in the continuous time model. The resulting three-parameter fit of the combined data resulted in the following fitted values (-/+ 95% confidence interval): post-assessment amplitudes 0.6039 (0.384, 0.8237) and 1.527 (1.457, 1.597), delta 1.092 (0.9544, 1.21), decay fraction 0.4099 (0.262, 0.5578). B) A Priori analysis of the number of days required to test whether two groups are significantly different. Power = 0.80 for the consensus average and standard deviation of the combined data from plasma ALT, AST, and CK for all participants.

To test the hypothesis that the temporal sequence of measurements is the origin of the significant changes observed in the features model, the analytes measured for each participant were randomly permuted in time. Bootstrap distributions and statistics (average and 95% bootstrap CI) were generated at each time point for all participant sequences with and without permutation (eFigure 4). All trends were removed following randomization of the time series data, indicating that model features are dependent upon the temporal ordering of the sequence. The correlations between physical assessment and morning activity with both the kinetic decay parameters and morning activity dependent offset (ΔZ-score) are significant observable features of the data.

To explore how temporal Z-score analysis may be useful in improving experimental design for interventional and longitudinal studies, a power analysis (Power = 0.80), based on the statistical properties of the observed data, was performed to address the number of samples required to identify 100% and 50% increases or decreases in ΔZ-score evaluated from an individual’s two sequences of blood collection, for three different probabilities of Type 1 error (α = 0.001, 0.01, and 0.05). Statistical significance is achievable within a time scale of approximately one month under this hypothesis. It remains to be tested if, with sufficient time series data before and after intervention, the ΔZ-score can be used to monitor progression and/or treatment efficacy. For example, a 50% decrease in ΔZ-score may be detected in 15 – 36 days if this participant cohort is representative of the population.

## DISCUSSION

While a single yearly measurement of creatine kinase (CK) cannot capture daily changes in the degree of sarcolemma fragility in FSMD once diagnosed, this study demonstrates that the kinetics of plasma ALT, AST, and CK can reproducibly mark sarcolemma fragility on time scales much shorter than a year: time-series analysis with appropriate normalization of plasma CK, ALT and AST correlated with activity across individuals and different participant populations monitored for one week. Activity-dependent changes were observed for all four enzymes, but their kinetics were not the same. ALT, AST, and CK behaved similarly: the feature model-derived lines before (blue) and after (red) morning activities were parallel as changes in the Z-scores were, on average, equal over the duration of the visit, whereas LDH’s lines converge (Figure 3). The change in Z-score was the same for ALT, AST, and CK, regardless of the baseline (blue) level before morning activity. This reproducibility in Z-score changes was remarkable, given that each participant arrived at the Clinical Center with a different baseline (elevated compared to subsequent measurements) and the choice and intensity of their morning activities were *ad libitum* for four continuous days. In addition, the decay in enzyme concentrations with bedrest each day was also reproducible and similar between participants. Thus, the consensus model generated from this analysis provides a parametrization of plasma ALT, AST, and CK kinetics that can guide future pathophysiological studies of FSMD.

It’s noteworthy that the blood draws in this study were irregularly spaced in time to capture both short (minutes, hours) and long (days) time periods correlated with activity permitting approximations of the underlying short and long-term kinetic behaviors. The high-frequency blood draws were designed to reveal the kinetics following routine physical assessment, while the data at other times were sparsely sampled to assess longer term differences. Since the total volume of blood drawn across the entire visit is constrained, the 30 minutes from bedrest through morning activity was not sampled and, consequently, is represented as a rapid linear increase or spike in the model (Figure 4). Regardless, the data demonstrate that there are significant positive correlations between physical activity and the kinetics of ALT, AST, CK, and LDH. The clearance rates calculated here for CK are also consistent with other reports, and plasma ALT and AST have previously been shown to scale with activity in muscular dystrophy and heat exhaustion-related rhabdomyolitis ^24-26^. Together, they demonstrate that the transformation of the data described here reflects the overall appearance into and clearance from the blood, which normally includes release from muscle, diffusion across the basement membrane, transfer to lymphatics, and passage to blood. Based on previous work ^27^, circadian rhythm is unlikely to significantly contribute to the temporal patterns observed. For individuals with FMSD, there is consensus that activity causes skeletal muscle enzymes to be released into the plasma, presumably due to skeletal muscle damage, that are subsequently cleared. Clearance can be studied using the methodologies developed here.

Unlike CK, ALT, and AST, the response of plasma LDH to the same morning activity decreased throughout the week, despite the average percent release of the four enzymes being similar (see below). This was curious and may be related to enhanced release of LDH early in the week, but not at the end of the week. Perhaps other activities prior to and during Days 1 and 2 (travel and physical assessments) sensitizes the LDH response to morning activities for those days. While almost every tissue has LDH, with heart, liver, and red blood cells obvious candidates for extra-skeletal muscle sources, there is no obvious reason for activity-dependent release of LDH from these other tissues. More work is needed to clarify this issue, including a study of the LDH isoenzyme composition.

### Time series analysis of plasma enzymes in diagnosis

Elevated plasma CK (20-100x normal levels) in an individual with muscle weakness currently serves as a diagnostic indicator, but not a definitive diagnosis, of muscular dystrophy ^28^ or necrotic inflammatory myopathy. This is partially due to the significant individual and interpersonal variability in plasma CK. This variation can be attributed to nearly every differentiating parameter, including sex, ancestry, ethnicity, age, physical condition, and even climate ^29-32^. Here, significant differences in plasma ALT, AST, CK, and LDH between LGMD R2 DYSF-related and R12 ANO-5 participant populations were found (Figure 2A and eFigure 1). This result expands upon previous findings that different muscular dystrophy subtypes (Becker’s, Duchenne, Emery-Dreifuss, facioscapulohumeral, and limb-girdle muscular dystrophies) have distinct serum enzyme profiles ^20, 33^. Since these differences in routine clinical chemistry analytes have broad overlap, even with the treatable necrotic inflammatory myopathies, genetic testing is still important to pursue. Whether sequence specific differences (genotype) within a specific LGMD subtype correlate with the magnitudes and time courses of activity dependent changes remains an open question.

Our analysis of plasma ALT, AST, CK and LDH supports and expands on findings that both inter- and intra-personal variation are important factors in population studies ^34^. By subtracting “baseline” concentrations and expressing all changes as variations around “baseline” before and after activity, significant variations in scaling between individuals are removed and correlated features are revealed. These temporal and correlated features would otherwise be lost when raw data is averaged because distributional properties and disparate scaling are not addressed.

### Sarcolemma damage in exercise

As in FSMD, inter- and intra-personal plasma CK variability also confound interpretation in exercise physiology. Plasma CK correlates with training intensity, maximum isometric strength, eccentric exercise, and muscle soreness ^29, 35^. Athletes can be classified as “high CK” and “low CK” responders based on exercise induced changes from baseline concentrations ^36, 37^. Without normalization, a single measurement of plasma CK in high responders can be misinterpreted as hyperCKemia or overexertion, while overexertion in low responders may be missed. Monitoring normalized plasma ALT, AST, and CK kinetics may help to maximize an individual’s training efficiency while avoiding overtraining ^37, 38^. Short-term, high-frequency blood draws during rigorous training would permit monitoring of enzyme kinetics and may provide a quantifiable indicator of how hard athletes should push themselves. For example, the detection of small changes in CK concentrations in low responders may allow earlier detection of rhabdomyolysis; early detection and intervention are essential for limiting or preventing loss of muscle mass, metabolic abnormalities, and acute kidney injury ^39, 40^.

Pathway analysis of biomarkers identified using high throughput biological data that correlate with physical activity may reveal their source; analytes whose concentrations increase with activity may originate from the same tissue or compartment, and analytes with similar decay times may enter the blood via similar mechanisms. Here, all four enzymes exhibit similar significant increases with activity (average % change +/- SEM across all participants with morning activity was ALT [12.8 ± 2.8%], AST [11.6 ± 2.9%], CK [12.9 ± 2.8%], LDH [12.2 ± 3.9%]), suggesting that they all originate from the same stimulated source, i.e., skeletal muscle. Prior studies have demonstrated that these enzymes may be released by damaged muscle ^41-43^. The mechanism(s) by which the enzymes enter the blood may combine leakage through sarcolemma wounds, exocytosis, extracellular vesicles ^44-46^ and activity-dependent lymph flow ^47, 48^.

One strength of this study is that our analysis permits investigations of activity-dependent correlations with plasma analytes independent of initial scaling and within small populations of individuals. A limitation is that our time series analysis was based on a small number of irregularly sampled time points. Longer time series are expected to improve the parameters of the feature model such that they are usable in time series clustering algorithms when comparing multiple plasma analytes. An additional weakness is specificity; CK-MM, the skeletal muscle specific isozyme was not assayed. While there were no indications of acute myocardial infarction or brain damage that could lead to increases in CK-MB or CK-BB, we cannot exclude that damage to these organs contributed to the reported findings. Future studies with higher time resolution and increased number of plasma analytes may increase the generalizability of these findings.

**In summary**, this study demonstrates that small populations can provide data that may benefit patient outcome if adapted to provide guidance on duration and activity minimizing muscle damage. As surrogate biomarkers of FSMD muscle membrane fragility, time- and activity-dependent measurements may aid diagnosis, long-term longitudinal studies of disease progression and efficacy of gene and drug therapies. The temporal properties of plasma ALT, AST, and CK may facilitate identification of biomarkers with similar, or different, kinetics that further elucidate sarcolemma damage and repair processes. Pathway analysis after classification of biomarkers by temporal features may reveal the origins and mechanisms by which these biomarkers enter the plasma, an approach that can be of utility in other diseases with different biomarkers.

## Supplementary Information

**eFigure 1:**
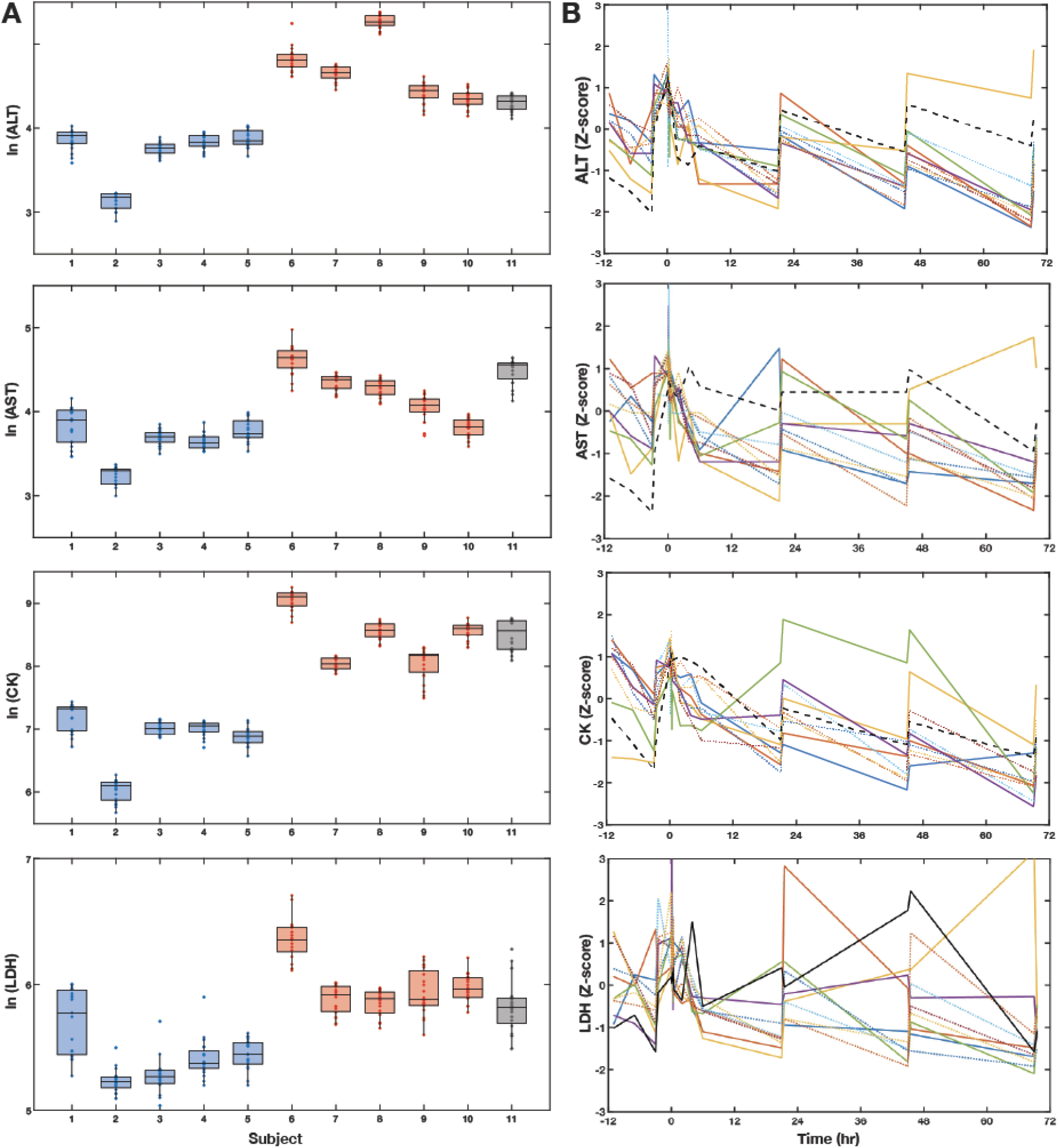
Transformed analysis of plasma ALT, AST, CK and LDH. A) Box plots of log transformed plasma ALT, AST, CK, and LDH by participant clustered by LGMD subtype (blue-R12 Anoctamin5-related; red-R2 Dysferlin-related; black- R9 FKRP-related). B) Z-transformed plasma enzyme levels for each participant over the study time course.

**eFigure 2:**
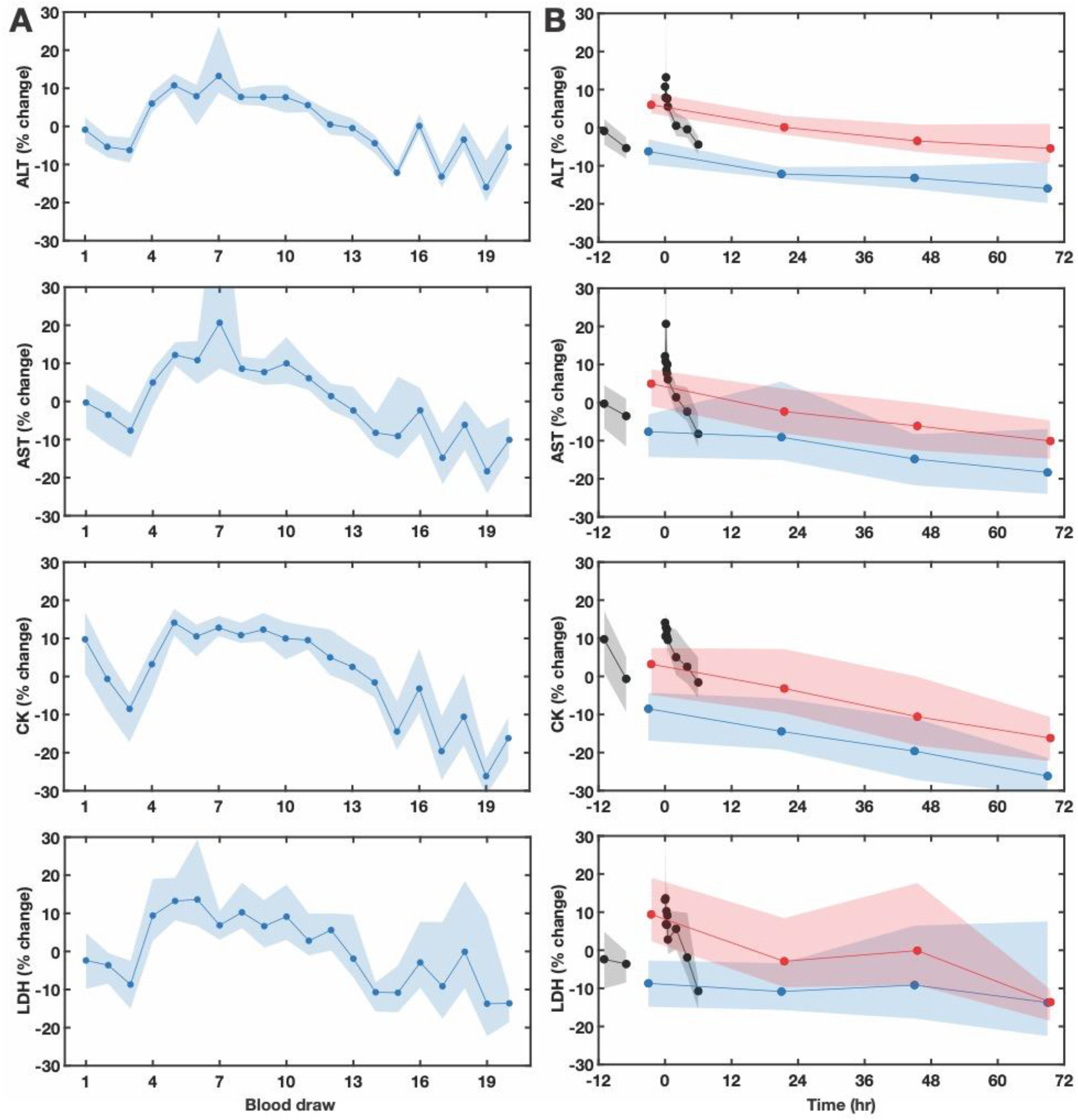
Percent change of ALT, AST, CK and LDH around the average follow similar trends to Z-transformed data as a function of blood draw and time. Percent change around the average (over all draws) by A) blood draw, and B) by time. Calculated as (value – average)/average. Average (connected points) and 95% bootstrap confidence intervals (shading).

**eFigure 3:**
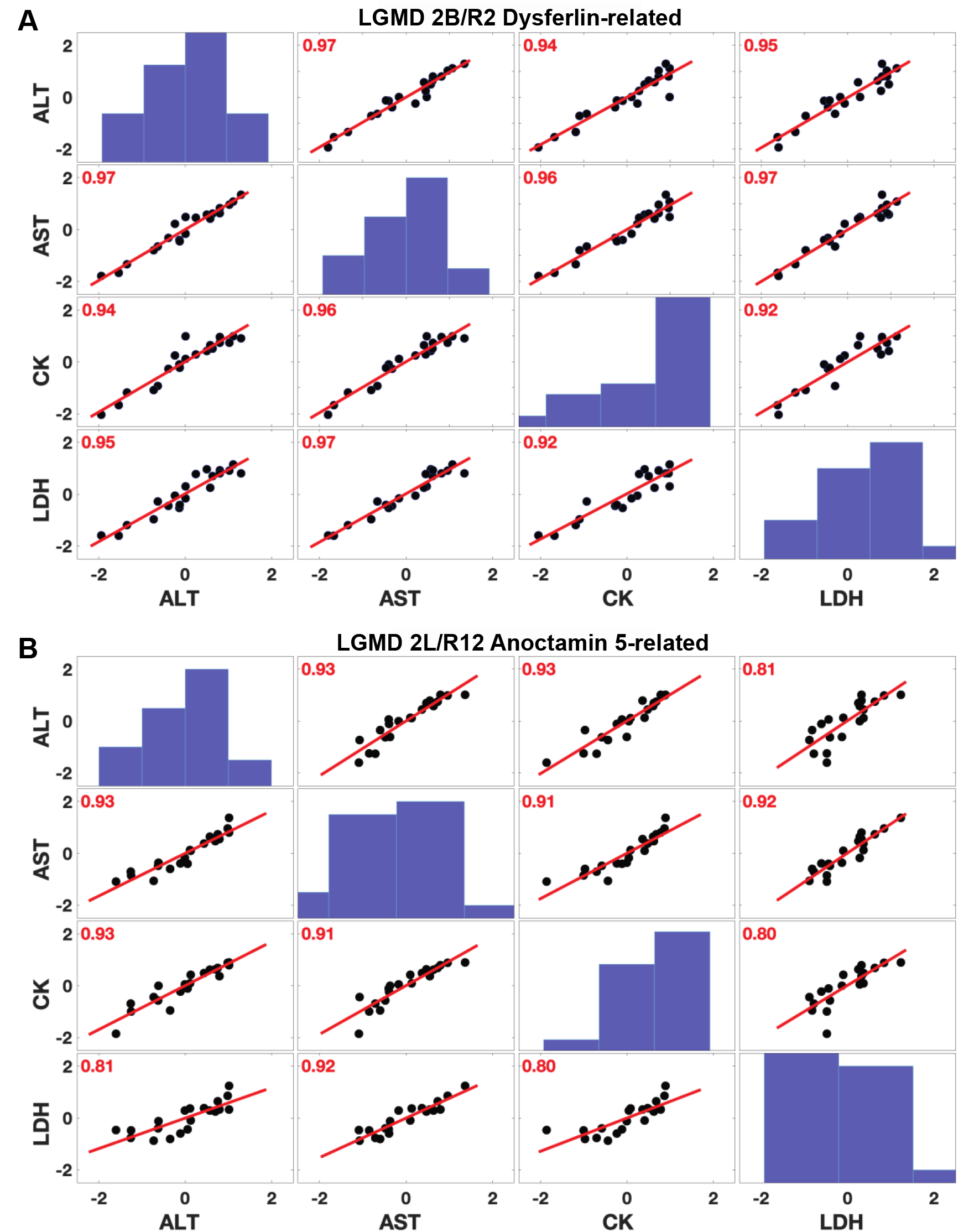
Pearson correlation analysis of ALT, AST, CK and LDH by LGMD subtype. A) LGMD 2B: R2 Dysferlin-related; B) LGMD 2L: R12 Anoctamin 5-related. The distributions of data are represented as histograms along the diagonal. All correlations are significant at the p = 0.05 level.

**eFigure 4:**
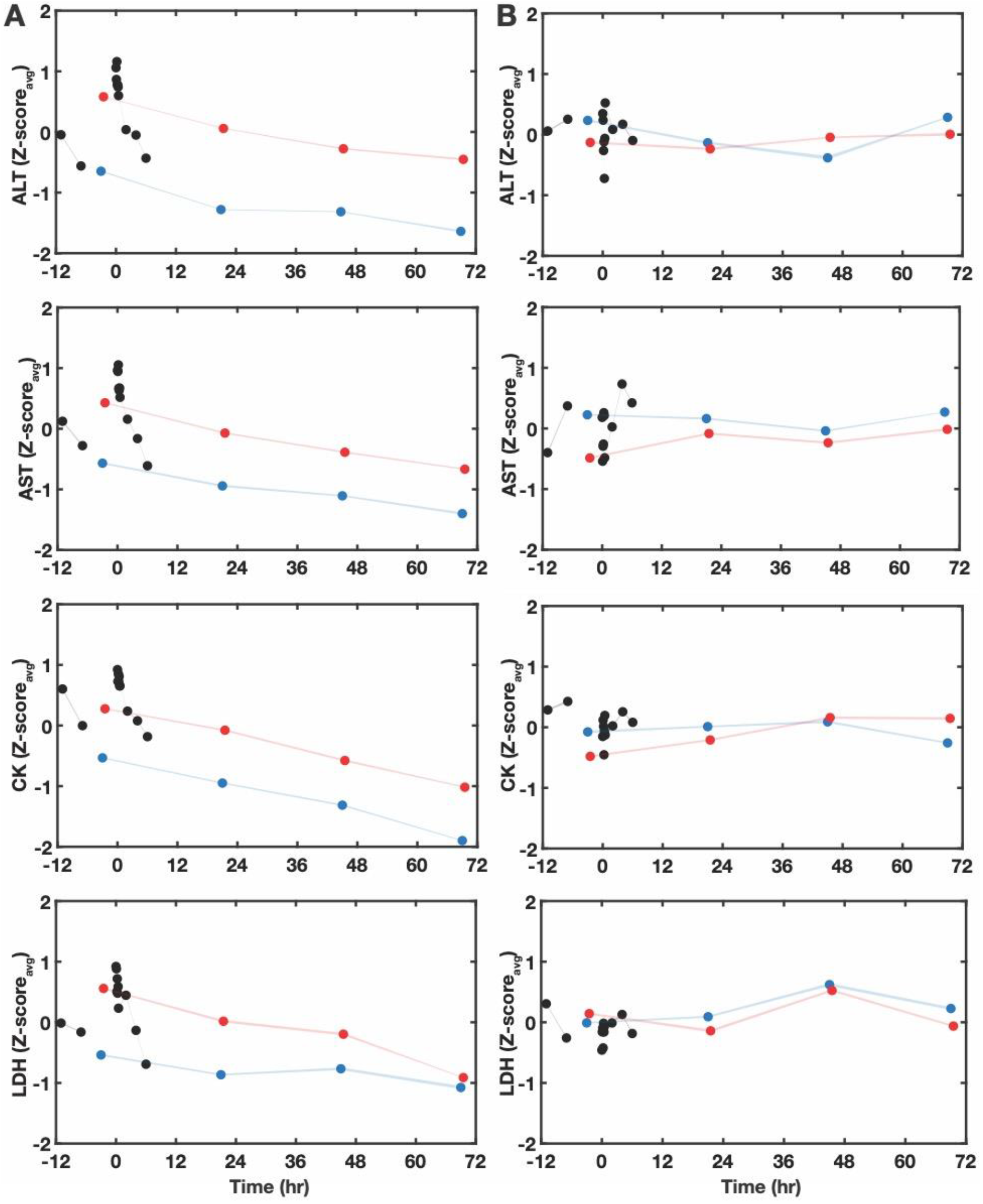
Average Z-score of plasma enzymes with random permutations in time. Average participant plasma ALT, AST, CK, and LDH Z-scores, without (A) and with (B) random permutations in time, plotted against time (in hours) with physical assessment at time 0. Blue and red timepoints correspond to pre- and post-morning activity; black points correspond to post-arrival and post-physical assessment.

**eTable 1:** A simplified model was used to fit the log transformed data, with one time constant for the high-frequency data, as the full feature model used for the Z-score data did not converge when applied to the log transformed data.

| Feature model | Fitted Values (-,+ 95% CI) |  |  |  |
| --- | --- | --- | --- | --- |
|  | ALT | AST | CK | LDH |
| Tau 2 (hr) | 41.82 (29.8, 53.85) | 31.3 (23.44, 39.16) | 34.59 (24.36, 44.82) | 36.87 (18.82, 54.92) |
| Tau long (hr) | 691.5 (398.7, 984.2) | 518.6 (326.4, 710.8) | 344 (253.8, 434.2) | 362 (142.7, 581.2) |

## eMethods

### Feature and continuous time models

A) Feature model: *I*_*1*_, *I*_*2*_, and *I*_*3*_ are linear intercepts, the linear slopes τ_1_, τ_2_, and τ_long_, correspond to exponential decay times, and *t*_*thresh*_ is the time point separating the two piecewise, linear components of the double exponential decay; B) Continuous time model: amplitudes Amp_1_ and Amp_2_, DZ, and fraction f_1_ are fitting parameters and all the τ_s_ were fixed at the values obtained using the feature model. The fractions f_1_ and (1-f_1_) represent the relative contribution of the processes with exponential decay τ_1_ and τ_2_, respectively.

A) Feature Model:

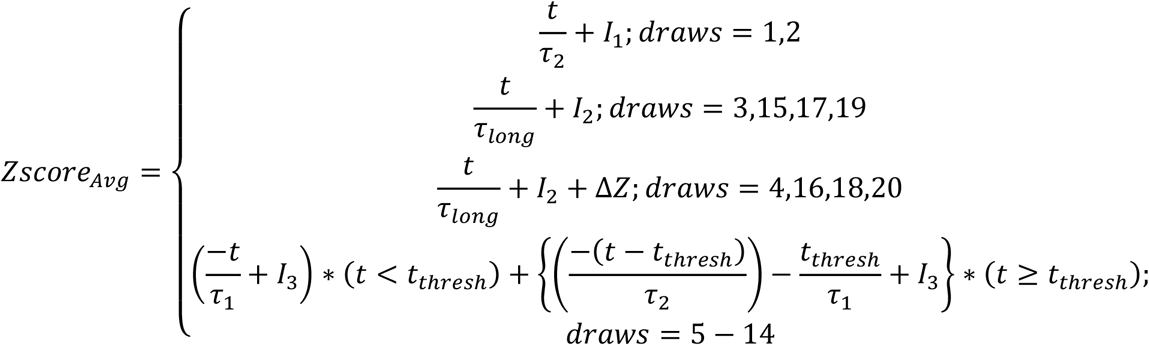

B) Continuous Time Model:

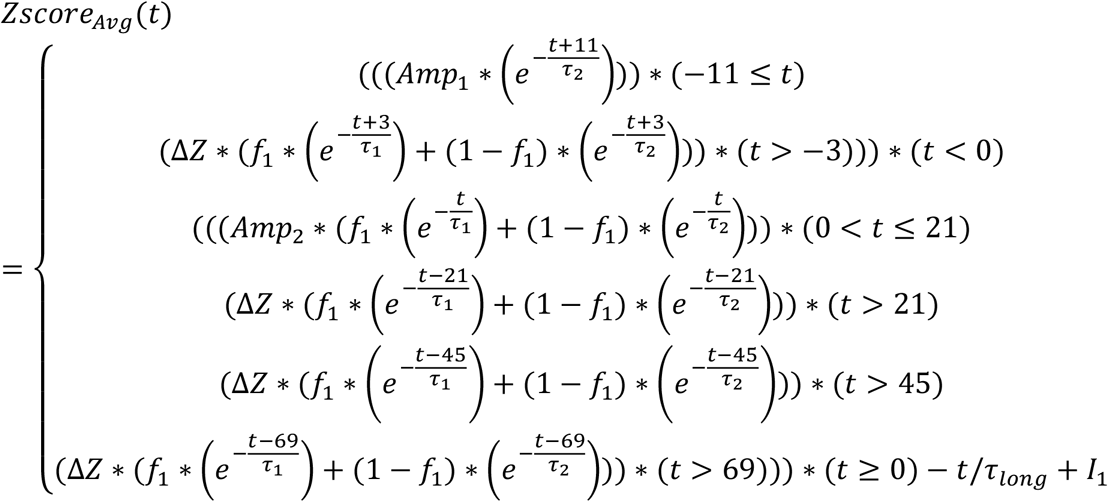

## Acknowledgments

The authors thank Ms. Irene T. Rozga, R.N., Dr. Gladys Tataw-Ayuketah, PhD, MPH, RN, and the staff of the Metabolic Clinical Research Unit team for their guidance and proficiency. The authors thank the Jain Foundation for their support of previous pre-clinical research that introduced us to the need for identifying surrogate biomarkers. The authors gratefully acknowledge the contributions of the study participants.

